# SARS-CoV-2 Detection from the Built Environment and Wastewater and Its Use for Hospital Surveillance

**DOI:** 10.1101/2021.04.09.21255159

**Authors:** Aaron Hinz, Lydia Xing, Evgueni Doukhanine, Laura A. Hug, Rees Kassen, Banu Ormeci, Richard J. Kibbee, Alex Wong, Derek MacFadden, Caroline Nott

## Abstract

**Background:** SARS-CoV-2 causes significant morbidity and mortality in health care settings. Our understanding of the distribution of this virus in the built healthcare environment and wastewater, and relationship to disease burden, is limited.

**Methods:** We performed a prospective multi-center study of environmental sampling of SARS-CoV-2 from hospital surfaces and wastewater and evaluated their relationships with regional and hospital COVID-19 burden. We developed and validated a qPCR-based approach to surface sampling, and swab samples were collected weekly from different locations and surfaces across two tertiary care hospital campuses for a 10-week period during the pandemic, along with wastewater samples.

**Results:** Over a 10-week period, 963 swab samples were collected and analyzed. We found 61 (6%) swabs positive for SARS-CoV-2, with the majority of these (n=51) originating from floor samples. Wards that actively managed patients with COVID-19 had the highest frequency of positive samples (p<0.01). Detection frequency in built environment swabs was significantly associated with active cases in the hospital throughout the study (p<0.025). Wastewater viral signal changes appeared to predate change in case burden.

**Conclusions:** Environment sampling for SARS-CoV-2, in particular from floors, may offer a unique and resolved approach to surveillance of COVID-19.

## BACKGROUND

COVID-19 has caused substantial morbidity and mortality worldwide since it was first identified at the end of 2019. Despite the development of effective vaccines and the discovery of multiple therapeutic agents^1,2^, the SARS-CoV-2 virus and agent of COVID-19 continues to transmit across all corners of the globe. As of Spring 2021, it is estimated that over 100 million people have been infected with SARS-CoV-2 from the start of the pandemic, with over 2.5 million deaths attributable to COVID-19^3^.

Hospitals were recognized early on as a potentially high-risk environment for acquiring COVID-19 infection for patients and healthcare workers, with healthcare facility outbreaks and nosocomial acquisition of COVID-19 common and well described^4^. While the respiratory droplet route is considered to be a primary means for transmission of SARS-CoV-2 in hospitals, it is still not well established if there is appreciable transmission through contact with the built environment^4,5^, and to what extent this could contribute to spread.

Laboratory experiments indicate that the virus can remain viable on stainless steel and plastics for approximately 6 hours and SARS-CoV-2 RNA can be detected from wastewater^6^. Preliminary studies have shown a range from no to extensive contamination of hospital ward surfaces, including computer mice, trashcans, and door handles^7^. In addition to identifying the potential routes for fomite-based transmission, surveying the distribution of SARS-CoV-2 throughout the hospital environment over time can provide insight into hospital burden and potentially outbreak identification and monitoring. New approaches for early outbreak detection will be particularly important to reduce institutional morbidity and mortality^8^. The built environment presence of SARS-CoV-2 could act as a spatially resolved indirect measure of symptomatic and asymptomatic carriage/infection within patients and healthcare workers (HCWs). In a similar fashion, wastewater surveillance may offer a less spatially resolved but potentially anticipatory mechanism for hospital COVID-19 surveillance and monitoring^9,10^.

In this study, we conducted a longitudinal assessment of the prevalence of SARS-CoV-2 on commonly contacted hospital surfaces as well as from hospital wastewater and compared these markers with active COVID-19 case prevalence at two large tertiary care hospitals and in the surrounding community.

## METHODS

### SARS-CoV-2 control samples

Control samples of heat-inactivated SARS-CoV-2 (ATCC VR-1986HK; lot 70035039) and SARS-CoV-2 RNA (ATCC VR-1986D; lot 70035624) were obtained from Cedarlane Laboratories. The copy numbers of the control samples were reported as 4.73 × 10^3^ genome copies/μL for the RNA and 3.75 × 10^5^ genome copies/μL for the heat-inactivated virus.

### Swabbing method and validation

Surfaces were sampled with P-208 Environmental Surface Collection Prototype kit from DNA Genotek, consisting of a flocked swab and a semi-lytic nucleic acid stabilization solution for post-collection swab immersion. The swabbing method was validated in laboratory experiments using surface materials spiked with heat-inactivated SARS-CoV-2. We applied 10^5^ copies of the virus to sheets of acrylic, stainless steel, and vinyl upholstery by pipetting 150 µl of virus suspension in 30-40 small spots within a 2.25 in by 2.25 in square. After a 2.5 hour drying period, swabs were pre-wetted with stabilization solution, surfaces were swabbed for 30 seconds, and swabs were stored in stabilization solution in collection vials. As controls, sterile swabs were placed in collection vials containing virus spiked directly into the stabilization solution. Following one day of storage at room temperature, RNA was extracted and analyzed by qPCR. The amount of virus in each sample was estimated using the virus standard curve that relates Cq values to input genome copies. The percent viral recovery was determined by dividing the amount of virus estimated for the surface swabbed samples by the amount of virus estimated in the control samples. Significant differences in recovery efficiencies across surfaces were determined by ANOVA with post hoc comparisons using Tukey’s honestly significant difference (HSD) test.

### RNA extractions and RT-qPCR

Presence of SARS-CoV-2 RNA was determined by quantitative reverse-transcriptase PCR (RT-qPCR) of RNA extracted from swab sample stabilization solution. RNA was extracted from samples using the MagMAX Viral/Pathogen II (MVP II) Nucleic Acid Isolation Kit (Thermo-Fisher Scientific) according to the manufacturer’s protocol, with reagent volumes modified proportionally for 300 µl sample input volumes. Extractions were performed in batches of up to 96 in deep-well plates. Each batch included positive (heat-killed SARS-CoV-2) and negative (stabilization solution only) control samples to monitor extraction efficacy and potential cross-contamination. None of the control samples yielded false positive or false negative results.

Quantitative PCR was performed with primers and a TaqMan probe targeting the N gene of SARS-CoV-2 (Table 1). The probe contained a 6-carboxyfluorescein (FAM) fluorescent dye at the 5’ end and the minor groove binder non-fluorescent quencher (MGB-NFQ) at the 3’ end. Each qPCR reaction consisted of 5 µl of extracted RNA, 300 nM of each primer, 200 nM of probe, and 5 µl of 4X TaqPath 1-Step Multiplex Master Mix (No ROX) (Applied Biosystems) in a final volume of 20 µl. Thermal cycling, fluorescence quantification, and quantitative cycle (Cq) determination were performed on a Bio-Rad CFX Connect with the following cycling conditions: 25 °C for 2 min, 50 °C for 15 min, 95 °C for 2 min, and 45 cycles of denaturation (95 °C for 3 s) and elongation (55 °C for 30 s).

**Table 1.**
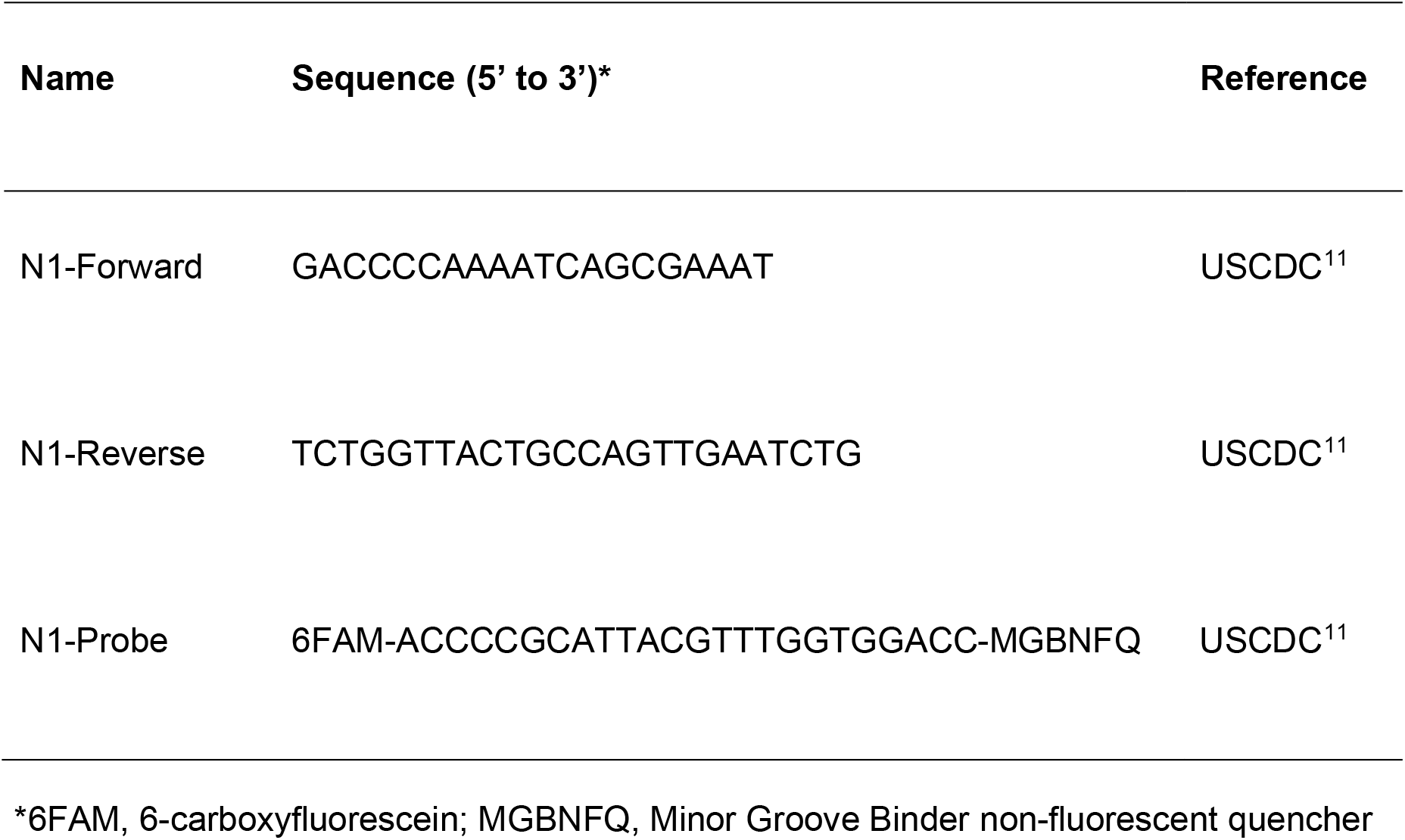
qPCR primer and probe sequences

### Estimates for Limits of Detection and Quantification

Analytical sensitivity and precision of the qPCR assays were determined by estimating the limits of detection and quantification of the SARS-CoV-2 RNA and virus standards. Serial 10-fold dilutions of the SARS-CoV-2 RNA standard were analyzed by qPCR with 14-20 replicates per dilution. For the virus standard, RNA was extracted from serial dilutions of heat-inactivated virus (7 replicate extractions at each dilution) prior to qPCR analysis.

The qPCR results were analyzed as described by Klymus et al.^12^ using the Limit of Detection (LOD) Calculator (R script available at: https://github.com/cmerkes/qPCR_LOD_Calc). The standard curves were modeled as linear regressions based on the middle two quartiles of Cq values for each standard concentration exhibiting >50% detection. The limit of detection was defined as the lowest input amount that was detected in >95% of replicates, and the limit of quantification was defined as the lowest input amount yielding reproducible Cq values with a coefficient of variation less than 0.35. For the RNA standards, the LOD with 95% confidence intervals and limit of quantitation (LOQ) were determined by a curve-fitting approach as described^12^. The virus LOD was estimated using a discrete threshold (*i*.*e*., the lowest tested input amount with >95% detection).

### Built Environment Sample Collection

We prospectively collected built environment swabs from two large tertiary-care, academic hospital campuses (Campus A and Campus B) from Ottawa, Ontario, Canada over a 10-week period between September 28 and December 6, 2020. We obtained between 40-50 samples from each campus weekly, generally maintaining location and surface type longitudinally, though we did not attempt to sample the exact same item longitudinally in order to improve our study generalizability. For example, a computer keyboard sampled in week 1 on Unit A, may be different from a computer keyboard sampled on Unit A in week 2. At both campuses, we collected samples from 1 COVID-dedicated unit, 1 ICU (COVID-designated area), 1 Emergency Department (ED) area, non-COVID medical and surgical units (n=3 at Campus A, n=4 at Campus B), 1 ambulatory unit (dialysis), public/non-patient-care spaces within the hospital (e.g. main elevators), and exterior hospital grounds (e.g. parking garage). A designated HCW space was also sampled at Campus A. Built environment samples were collected by wetting a P-208 kit flocked swab with the stabilization media, and sweeping the swab tip across an approximately 2” x 2” area (dependent upon the item/surface size) for approximately 30 seconds. The swab was then transported in the P-208 stabilization media, and stored at ambient temperature to be processed/analyzed within 1 month of collection. Three study authors (LX, DM, CN) collected the samples during the study period, and swab technique was harmonized between them at onset of sample collection. A run-in week was performed prior to week 1, in order to test sample collection, logistics, and sample processing, but is not included in this analysis as a number of specimen sites were initially not included for this week, making inter-week comparisons challenging.

Specific (non-critical care) wards in the hospital were designated for the ongoing inpatient care of COVID patients, and these were designated as ‘COVID Wards’. However, we classified both the COVID Wards as well as the ICU and ED as ‘COVID Risk Units’, because they were anticipated to be managing a large number of SARS-CoV-2 infected patients.

### Hospital Practices

The visitor policy at the start of the study period limited visitation to 1 visitor/patient/day; however, at the start of study week 4, visitors were further restricted to essential caregivers only. Environmental cleaning was in keeping with provincial best practice and occurred daily in both public and clinical areas, with all contact points disinfected with Vert-2-Go© everyday disinfectant (quaternary-ammonium based product) and floors mopped with Vert-2-Go© Oxy floor cleaner. The only deviation from this was once weekly cleaning of the Campus A parking garage until sometime between study weeks 5-7, then once daily thereafter. Infection Prevention and Control (IPAC) policies were also consistent with provincial best practice^6,13^. Throughout the study period, all staff, visitors, and patients were screened for COVID-19 symptoms and risk factors (e.g. exposures, international travel) on entry to the hospital, in accordance with Ontario Ministry of Health guidance^14^. Universal masking for all staff and visitors was introduced prior to the study period. Universal eye protection (visor or goggles) for staff working in clinical areas was introduced at the start of study week 1.

### Wastewater Sample Collection

Raw sewage grab samples from Campus A were taken with a 5L bucket on a rope between 11:10am and 12:30pm each sampling day, which occurred once every 1 to 2 weeks during (and immediately prior to) the study period. The samples were swirled in the bucket and quickly poured into pre-labelled sterile 1L Nalgene bottles which were immediately placed into a cooler with ice packs for transport back to the lab and processed within 1 hr. Temperature of the samples, measured at the time of sampling with an infrared thermometer, for Oct-Dec were between 12 and 22°C. A 200ml aliquot was concentrated to 0.25-1.25ml using the Centricon® Plus-70 (MilliporeSigma) centrifugal ultrafilter with a cut-off of 10 kDa^15^. The resulting concentrate was extracted using the RNeasy PowerMicrobiome Kit (QIAGEN) according to the manufacturer’s instructions with an elution of 100µl.

Primers and probes, and Real-Time Reverse Transcriptase (RT RT-PCR) methods developed by the US CDC were used with the Bio-Rad CFX96 Touch Real-Time PCR Detection System. The RT RT-PCR thermal cycling protocol used to amplify the target region within the SARS-CoV-2 nucleocapsid gene, N2, was carried out at 50 °C for 30 minutes, followed by 95 °C for 15 minutes and 50 cycles of 95 °C for 15 and 60 °C for 30 seconds. Reaction mixture for RT RT-PCR, 5 μl RNA template, 4 μl of QIAGEN OneStep RT-PCR 5x Buffer, 2 μl dNTP mix (10 mM each), 2 μl Enzyme Mix, Primers and Probes were used at 500nM/reaction, and 2 µl of 4 mg/ml BSA. Each reaction volume was adjusted to a final volume of 20 µl with RNase-free water. Each sample was run in duplicate. An MS-2 bacteriophage matrix spike whole process control was used to assess the efficiency of the concentration process and RNA extraction as well as confirm RT RT-PCR protocols. RT RT-PCR inhibition control using MS2 bacteriophage spiked into raw sewage RNA extracts and molecular grade water was used to assess the performance of the RNA-extraction, RT-PCR and to detect the presence of inhibitors. Quantification of the N2 gene target in the raw sewage sample concentrates was done using standard curves created with a 2019-nCoV_N_Positive Control plasmid (IDT, Inc.) Non-template controls (NTC) were run for each mastermix preparation with molecular grade water as the template. For each week under observation, we averaged the detectable copies/ml from both N1 and N2 gene targets.

### Hospital and Citywide COVID-19 Cases and Outbreaks

Daily active patient COVID-19 case counts and locations, including admitted and non-admitted Emergency Department cases, were obtained from Infection Prevention and Control (IPAC). Cases were those confirmed by RT-PCR. We generated a measure of weekly ‘active’ COVID-19 cases by taking the mean of these daily active cases for each week (typically data only available for weekdays). Healthcare worker (HCW) cases were not included in the analysis, as HCWs were more likely than patients to spend time on multiple units and move throughout the hospital. Also, once a HCW became symptomatic or had a positive COVID-19 test result, they would no longer remain within the hospital. Hospital outbreak data were obtained in real time from IPAC and verified post-study on the publicly available Ottawa Public Health (OPH) COVID-19 dashboard^16^. An outbreak was defined as per the Ontario Ministry of Health^17^ as, “Two or more laboratory-confirmed COVID-19 cases (patients and/or staff) within a specified area (unit/floor/service) within a 14-day period where both cases could have reasonably acquired their infection in the hospital”. New and total active COVID-19 cases in Ottawa were obtained daily from the OPH COVID-19 dashboard, and a weekly ‘active’ caseload was calculated as the mean of the daily active cases across a given week.

### Data Analysis

To evaluate the characteristics associated with positive swab results, we performed a linear regression model using specimen characteristics as predictors of built environment sample positivity (total or floor specific specimens). Specimen characteristics were coded as categorical variables, including unit COVID Risk type, object/site, material, study week. The outcome (sample positivity) was coded as a binary variable (1=detected, 0=not-detected). A linear model was used to predict the outcome variable as it provided improved interpretability of coefficients and also model convergence stability^18^.

We also evaluated the strength of associations between (i) proportion of hospital built environment swabs positive for SARS-CoV-2 and (ii) proportion of hospital floor swabs positive for SARS-CoV-2, and hospitalized active COVID-19 cases. We used a negative-binomial regression model with active cases as the outcome. We did not quantitatively evaluate the association between wastewater and active cases because of the limited number of wastewater data points.

For the majority of the analyses we analyzed the two campuses in combination, as they reflect a single health care center with transfer occurring within and between units in both campuses. We also chose not to evaluate case burden by specific unit because, in general, as soon as a case was identified on a given non-COVID risk unit, the patient would be transferred to a specific COVID Risk unit either for cohorting or due to severity of illness (e.g. ICU).

All statistical analyses were performed using R (Version 4.0.3). Research ethics board approval was obtained through the Ottawa Health Science Network Research Ethics Board (OHSN-REB) for this study.

## RESULTS

### Limit of Detection and Swab Validation

We detected and quantified SARS-CoV-2 RNA by qPCR analysis using the N1 primer and probe set targeting the nucleocapsid gene^11^. A standard curve relating the quantification cycle (C_q_) values to viral genome copy number was produced by analyzing a 10-fold dilution series of a SARS-CoV-2 RNA standard (Fig. 1A). We used a model-fitting approach to determine the LOD, defined as a detection rate greater than 95%, and LOQ defined as a C_q_ coefficient of variation less than 0.35^12^. Reactions containing at least 3 genome copies were reliably detected under our qPCR conditions, whereas precise quantification required at least 150 copies (C_q_ < 30) (Fig. 1A).

**Figure 1.**
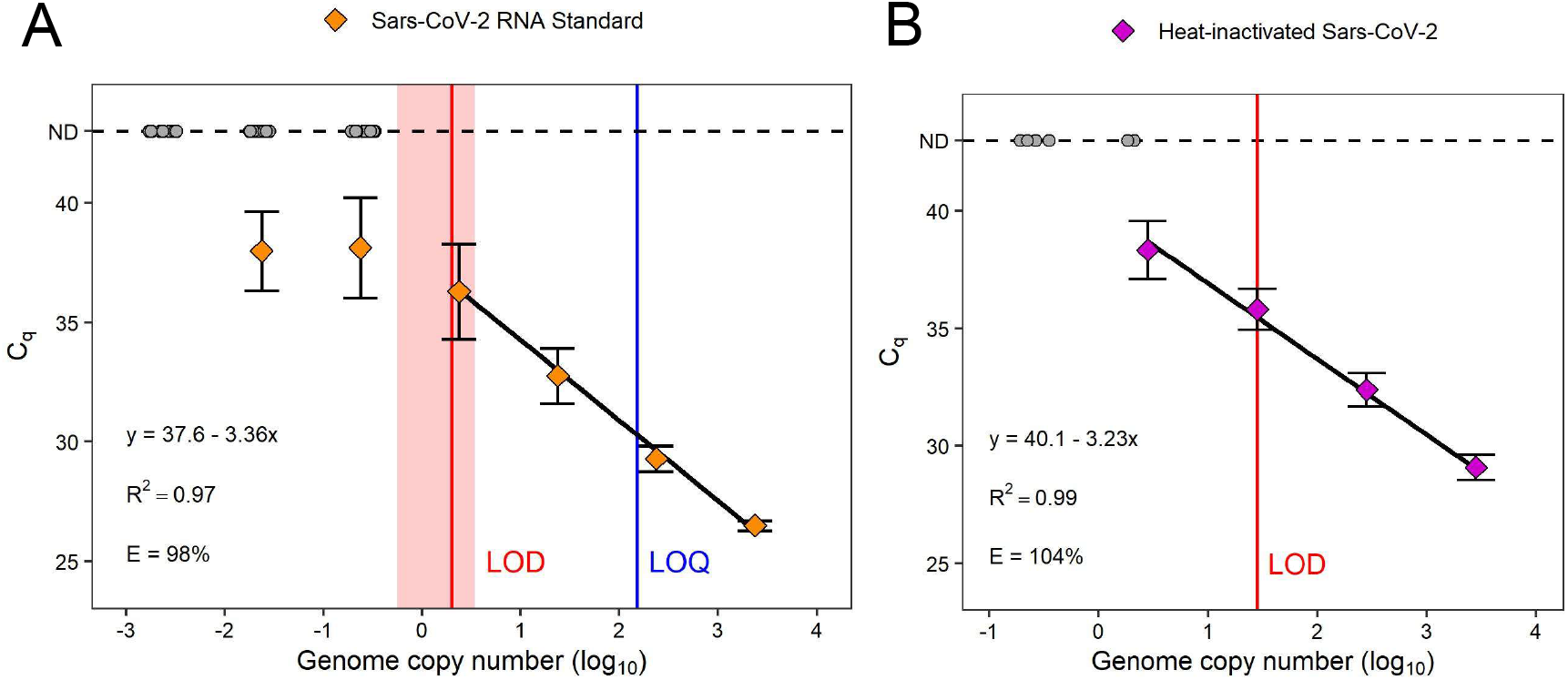
Limit of Detection analysis of SARS-CoV-2 RNA and virus standards. Serial dilutions of (A) SARS-CoV-2 RNA standard and (B) nucleic acid extractions of heat-inactivated virus were analyzed by qPCR targeting the N1 gene. Mean Cq values and standard deviations are plotted for 14-20 replicates (RNA standard) or 7 replicates (viral extractions). Grey circles indicate replicates that were not detected (ND). The limit of detection (red line with shaded 95% confidence interval) and limit of quantification (blue line) for the RNA standard were estimated by a model-fitting approach. The LOD for the viral extractions was estimated by a discrete threshold of >95% positivity. Equations, R^2^ values, and amplification efficiency (E) are shown for each standard curve.

Analysis of the built environment swab samples involved a nucleic acid extraction step prior to qPCR analysis. We evaluated the extraction method by qPCR analysis of RNA extracted from samples of a heat-inactivated SARS-CoV-2 virus standard (Fig. 1B). The LOD for the virus standard (28 copies per reaction) was ∼10-fold higher than that of the RNA standard, which can be due to RNA loss during the extraction procedure, presence of PCR inhibitors in the eluant, or inaccuracies in the reported titers of the standards. Nevertheless, the procedure was sensitive and reproducible, and the results yielded a linear standard curve for converting C_q_ values to viral copies while accounting for inaccuracies introduced by sample processing.

The built environment sampling approach relied on recovery of virus particles and nucleic acid from surfaces. In preliminary tests, we determined how efficiently our swabbing technique recovered virus from surface types representative of the built environment: acrylic, stainless steel, and vinyl. The surfaces were spiked with equal amounts of heat-inactivated virus, allowed to dry, and swabbed by the procedure used for built environment sampling. Following incubation of the swabs in stabilization solution and qPCR analysis of extracted RNA, we estimated the amount of recovered virus using the virus standard curve and calculated the percent of virus recovered from each surface (Fig. 2). The mean recovery efficiencies ranged from 28 to 42% across the three surfaces, values similar to those reported in a previous study of SARS-CoV-2 surface swabbing^19^.

**Figure 2.**
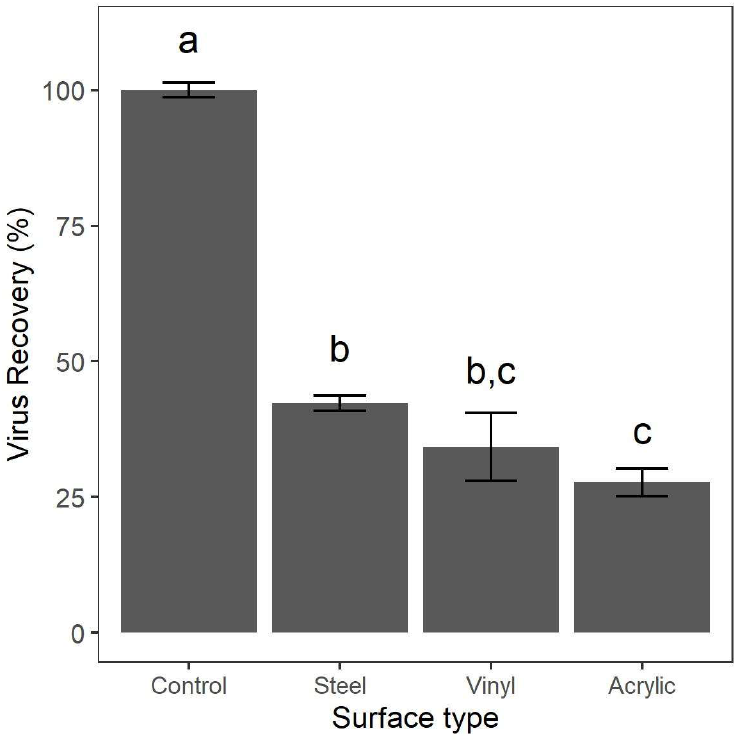
Swabbing recovery of heat-inactivated SARS-CoV-2 virus from spiked surfaces. Test surfaces were spotted with 10^5^ copies of the virus standard, allowed to dry, and swabbed for 30 seconds. Swabs were stored in nucleic acid stabilization solution overnight, RNA was extracted, and recovered genome copies were quantified by qPCR analysis. Virus recovery was determined by dividing the copies estimated for each sample by the copies estimated for a “no swabbing” control. The mean and standard error for three replicates is shown. Letters indicate significant differences between groups, determined by one-way ANOVA followed by Tukey’s post hoc test.

### Built Environment Screening

Over a 10-week period from September 28th to December 6th 2020 we systematically performed 963 built environment swabs across 2 tertiary care hospital sites. SARS-CoV-2 was detected from 6% of swabs (61/963), and was most commonly found from floor samples, with a detection prevalence of 27% (51/188). SARS-CoV-2 was less frequently identified from elevator buttons, benches, items (*e*.*g*., hand sanitizer pumps), and computer keyboards.

COVID Risk units, namely COVID treatment wards, intensive care units, and emergency rooms, were the most common locations to find SARS-CoV-2 in the hospital environment, with detection prevalence of 18%, 8%, and 7% respectively (Table 2, Fig. 3). Non-COVID and public/HCW spaces tended to have a low frequency of SARS-CoV-2 detection from the environment. During the course of this study, there were 2 outbreaks involving study units, at Campus B in the ED (1 patient and 2 HCW) and at Campus A on Ward 3 (5 patients and 1 HCW). A third outbreak occurred on a non-COVID inpatient unit at Campus A but this unit was not sampled in the study.

**Table 2.** Built environment swab SARS-CoV-2 detection by location and characteristics.

|  | Campus A |  | Campus B |  | Total |  |
| --- | --- | --- | --- | --- | --- | --- |
|  | Detected<br>(N=39) | Not-Detected<br>(N=451) | Detected<br>(N=22) | Not-Detected<br>(N=451) | Detected<br>(N=61) | Not-Detected<br>(N=902) |
| **Unit COVID Risk Type** |  |  |  |  |  |  |
| COVID Risk Unit | 17 (43.6%) | 150 (33.3%) | 17 (77.3%) | 126 (27.9%) | 34 (55.7%) | 276 (30.6%) |
| Non-COVID Unit | 9 (23.1%) | 221 (49.0%) | 5 (22.7%) | 266 (59.0%) | 14 (23.0%) | 487 (54.0%) |
| Public/HCW Area | 13 (33.3%) | 80 (17.7%) | 0 (0%) | 59 (13.1%) | 13 (21.3%) | 139 (15.4%) |
| **Object/Site** |  |  |  |  |  |  |
| Air (Control) | 0 (0%) | 10 (2.2%) | 0 (0%) | 10 (2.2%) | 0 (0%) | 20 (2.2%) |
| Computer Keyboard | 0 (0%) | 75 (16.6%) | 1 (4.5%) | 77 (17.1%) | 1 (1.6%) | 152 (16.9%) |
| Door Handle | 0 (0%) | 88 (19.5%) | 0 (0%) | 97 (21.5%) | 0 (0%) | 185 (20.5%) |
| Elevator Buttons | 4 (10.3%) | 46 (10.2%) | 2 (9.1%) | 57 (12.6%) | 6 (9.8%) | 103 (11.4%) |
| Exterior Bench | 1 (2.6%) | 9 (2.0%) | 0 (0%) | 0 (0%) | 1 (1.6%) | 9 (1.0%) |
| Floor | 32 (82.1%) | 59 (13.1%) | 19 (86.4%) | 78 (17.3%) | 51 (83.6%) | 137 (15.2%) |
| Hand Sanitizer | 1 (2.6%) | 49 (10.9%) | 0 (0%) | 59 (13.1%) | 1 (1.6%) | 108 (12.0%) |
| Other Item | 1 (2.6%) | 37 (8.2%) | 0 (0%) | 10 (2.2%) | 1 (1.6%) | 47 (5.2%) |
| Table | 0 (0%) | 3 (0.7%) | 0 (0%) | 2 (0.4%) | 0 (0%) | 5 (0.6%) |
| Telephone Receiver | 0 (0%) | 75 (16.6%) | 0 (0%) | 61 (13.5%) | 0 (0%) | 136 (15.1%) |
| **Material** |  |  |  |  |  |  |
| Air (Control) | 0 (0%) | 10 (2.2%) | 0 (0%) | 9 (2.0%) | 0 (0%) | 19 (2.1%) |
| Metal | 2 (5.1%) | 120 (26.6%) | 2 (9.1%) | 154 (34.1%) | 4 (6.6%) | 274 (30.4%) |
| Plastic/Vinyl | 37 (94.9%) | 318 (70.5%) | 20 (90.9%) | 287 (63.6%) | 57 (93.4%) | 605 (67.1%) |
| Wood | 0 (0%) | 3 (0.7%) | 0 (0%) | 1 (0.2%) | 0 (0%) | 4 (0.4%) |
| **Week** |  |  |  |  |  |  |
| 1 | 7 (17.9%) | 41 (9.1%) | 4 (18.2%) | 44 (9.8%) | 11 (18.0%) | 85 (9.4%) |
| 2 | 6 (15.4%) | 43 (9.5%) | 2 (9.1%) | 45 (10.0%) | 8 (13.1%) | 88 (9.8%) |
| 3 | 6 (15.4%) | 40 (8.9%) | 3 (13.6%) | 45 (10.0%) | 9 (14.8%) | 85 (9.4%) |
| 4 | 8 (20.5%) | 39 (8.6%) | 2 (9.1%) | 46 (10.2%) | 10 (16.4%) | 85 (9.4%) |
| 5 | 3 (7.7%) | 47 (10.4%) | 3 (13.6%) | 43 (9.5%) | 6 (9.8%) | 90 (10.0%) |
| 6 | 3 (7.7%) | 47 (10.4%) | 2 (9.1%) | 44 (9.8%) | 5 (8.2%) | 91 (10.1%) |
| 7 | 3 (7.7%) | 47 (10.4%) | 3 (13.6%) | 45 (10.0%) | 6 (9.8%) | 92 (10.2%) |
| 8 | 2 (5.1%) | 48 (10.6%) | 1 (4.5%) | 46 (10.2%) | 3 (4.9%) | 94 (10.4%) |
| 9 | 1 (2.6%) | 49 (10.9%) | 0 (0%) | 47 (10.4%) | 1 (1.6%) | 96 (10.6%) |
| 10 | 0 (0%) | 50 (11.1%) | 2 (9.1%) | 46 (10.2%) | 2 (3.3%) | 96 (10.6%) |

**Figure 3.**
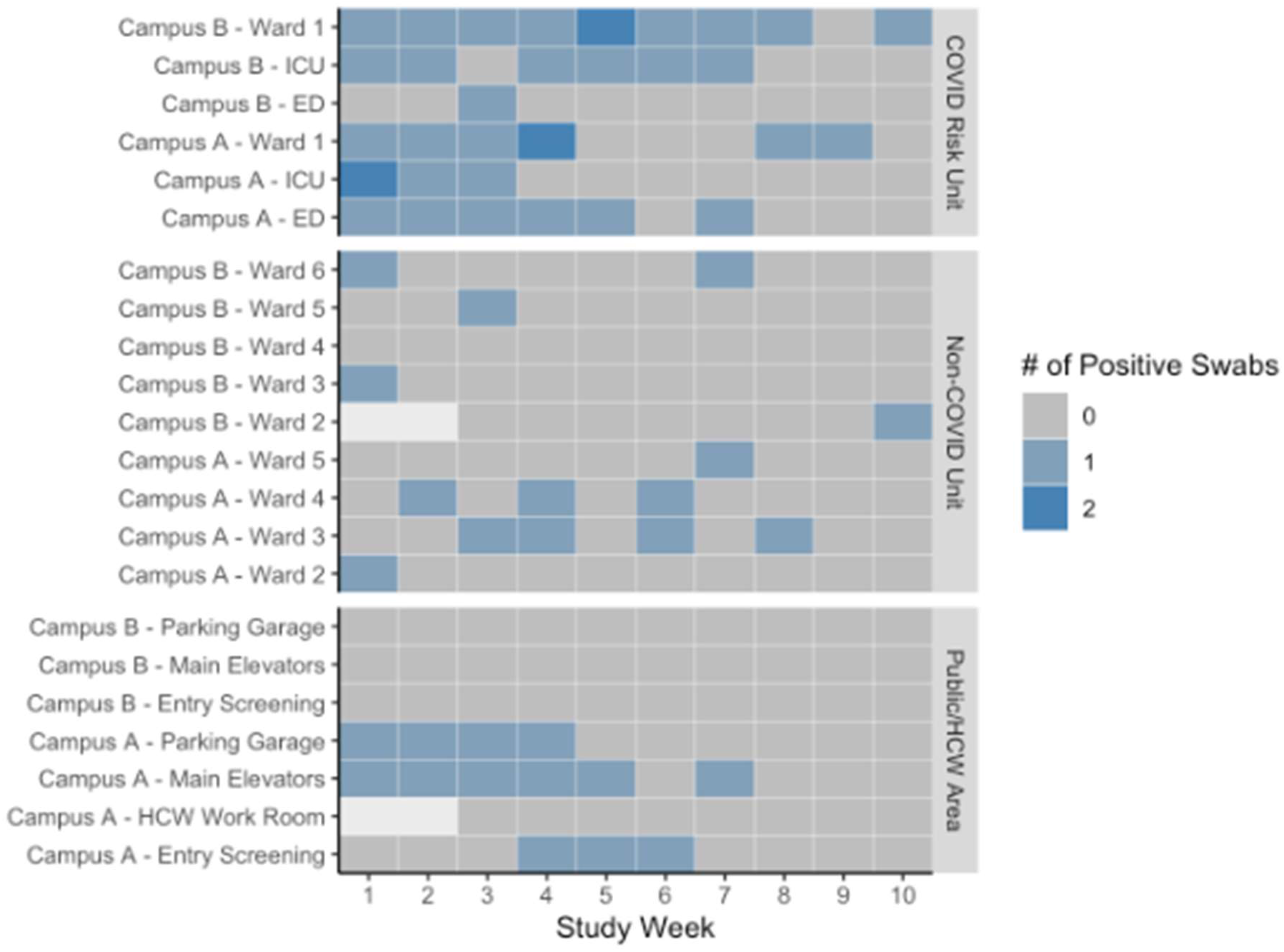
Time series of built environment swabs positive for SARS-CoV-2 by COVID risk location. *\*Blank (white) cells represent weeks where no swab was collected for a given location*.

Across the 10-week study period, SARS-CoV-2 detection prevalence paralleled total active cases across the two hospital campuses. Viral prevalence gradually declined over time, but with spikes in hospitalized cases being clearly reflected in built environment burden (Figure 4A), as well as an increase reflecting the start of hospitalizations in the second wave of the pandemic within the city. Built environment burden also paralleled citywide cases, but with less concordance (Figure 4B). Campus B tended to have SARS-CoV-2 positivity largely restricted to COVID Risk units, but Campus A seemed to have more activity in non-COVID risk units/public spaces. A multi-variable logistic regression model identified that increasing study week was associated significantly with reduced detection of SARS-CoV-2 (p<0.05), and that COVID Risk units and Campus A were significantly associated with increased SARS-CoV-2 detection (Table 3, Table 4).

**Figure 4.**
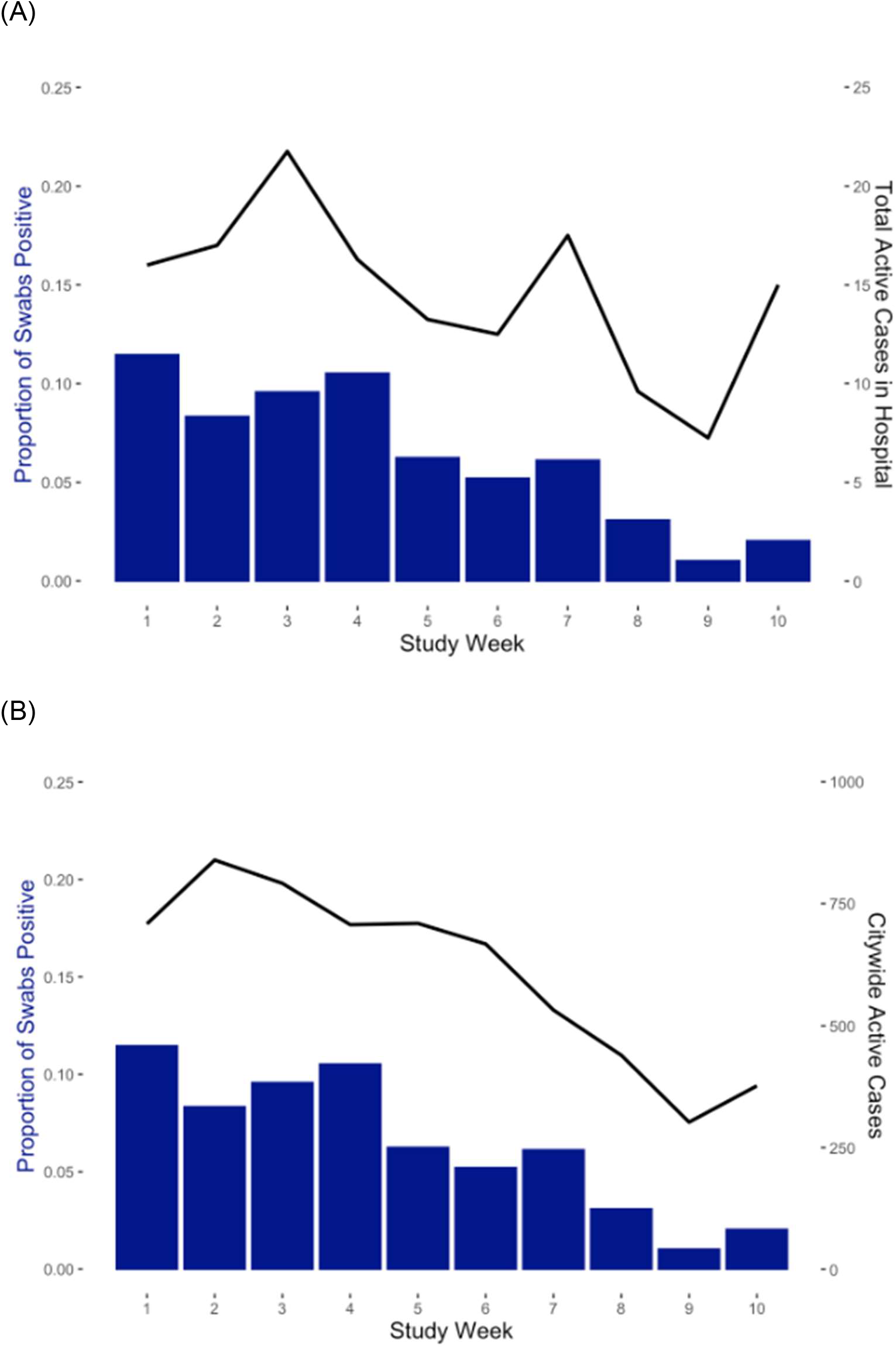
Proportion of hospital built environment swabs positive for SARS-CoV-2 (bars) and [A] mean hospital COVID-19 active cases (line) or [B] mean citywide COVID-19 active cases (line).

**Table 3.** Swab characteristics and measure of association with SARS-CoV-2 detection.

| Characteristic | Beta | 95% CI<sup>†</sup> | p-value |
| --- | --- | --- | --- |
| Study Week | -0.01 | -0.02, -0.01 | <0.001 |
| Material |  |  |  |
| Metal | — | — |  |
| Air (Control) | 0.12 | -0.32, 0.56 | 0.6 |
| Plastic/Vinyl | 0.07 | 0.00, 0.14 | 0.065 |
| Wood | -0.02 | -0.50, 0.46 | >0.9 |
| Specific Site |  |  |  |
| Air (Control) | — | — |  |
| Computer Keyboard | 0.05 | -0.38, 0.47 | 0.8 |
| Door Handle | 0.10 | -0.33, 0.53 | 0.6 |
| Elevator Buttons | 0.18 | -0.25, 0.61 | 0.4 |
| Exterior Bench | 0.20 | -0.25, 0.66 | 0.4 |
| Floor | 0.31 | -0.11, 0.74 | 0.15 |
| Hand Sanitizer | 0.05 | -0.37, 0.48 | 0.8 |
| Other Item | 0.05 | -0.38, 0.47 | 0.8 |
| Table | 0.05 | -0.55, 0.65 | 0.9 |
| Telephone Receiver | 0.04 | -0.39, 0.47 | 0.9 |
| Unit COVID Risk Type |  |  |  |
| Non-COVID Unit | — | — |  |
| COVID Risk Unit | 0.08 | 0.05, 0.11 | <0.001 |
| Public/HCW Area | 0.02 | -0.02, 0.06 | 0.3 |
| Campus |  |  |  |
| Campus B | — | — |  |
| Campus A | 0.03 | 0.00, 0.06 | 0.033 |

<sup>†</sup> CI = Confidence Interval

**Table 4.** Univariate coefficients with 95% confidence intervals (CI), p-values, and Akaike information criterion (AIC) for prediction of hospital case burden from (i) proportion of built environment swabs positive and (ii) proportion of floor swabs positive.

| Variable | Coefficient | 95% CI | P-value | AIC |
| --- | --- | --- | --- | --- |
| All Built Environment Swabs | 5.8 | 1-11 | 0.02 | 56.5 |
| Floor Swabs | 1.7 | 0.5-3.1 | 0.01 | 54.9 |

Hospital floors were the most common surface from which SARS-CoV-2 was detected. We looked specifically at prevalence on floors and its relationship with cases within the hospital and citywide (Figure 5). Restricting analysis to only floors showed they parallel the findings seen with all samples (Figures 4), with stronger correlation with active hospital cases.

**Figure 5.**
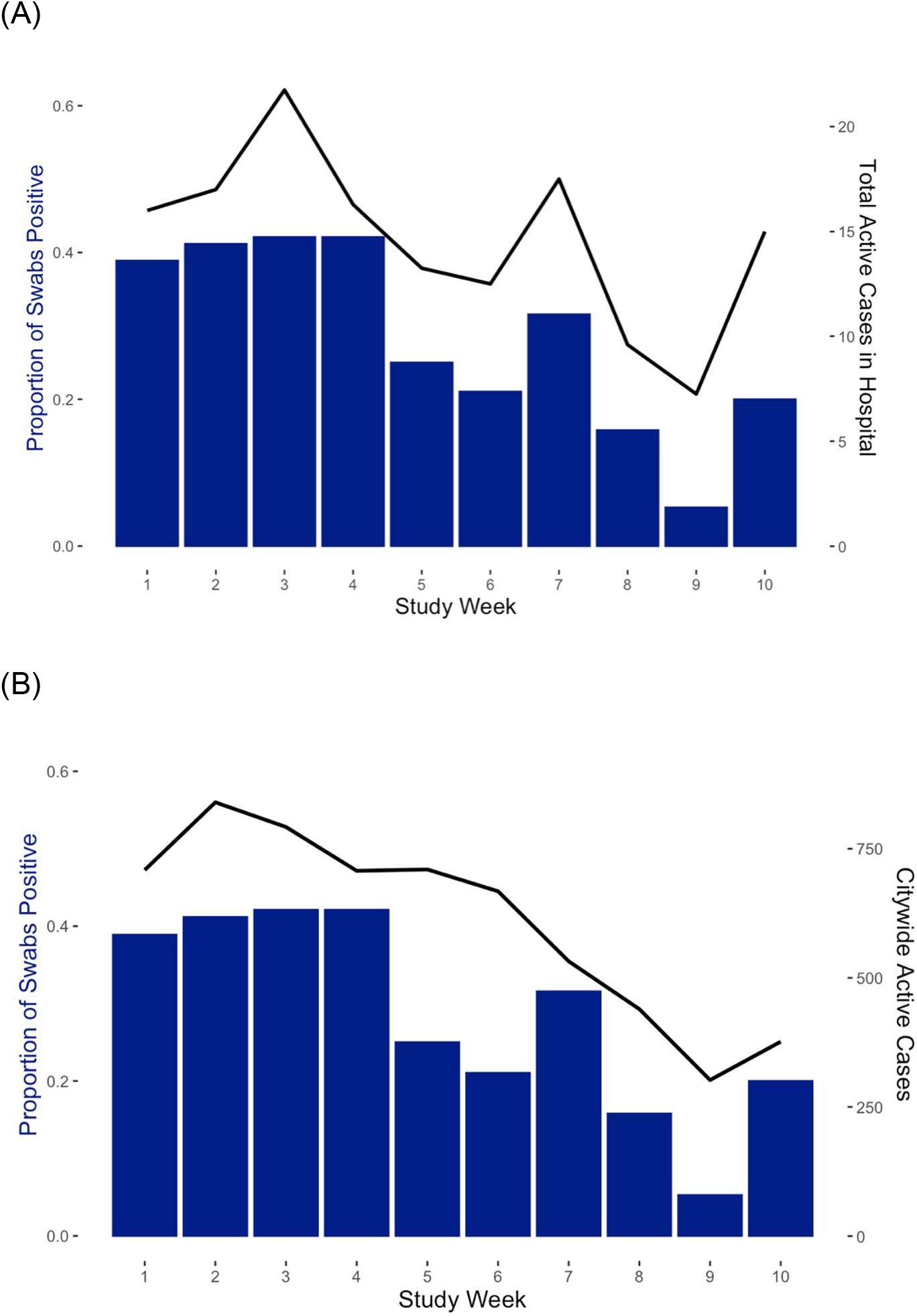
A. Proportion of hospital floor swabs positive for SARS-CoV-2 (bars) and [A] mean hospital COVID-19 active cases (line) or [B] mean citywide COVID-19 active cases (line).

Hospital wastewater SARS-CoV-2 viral copies per ml (Figure 6) followed a similar trajectory as built environment sampling but decline in wastewater levels appeared to predate both the built environment sampling as well as Campus A and citywide case burden.

**Figure 6.**
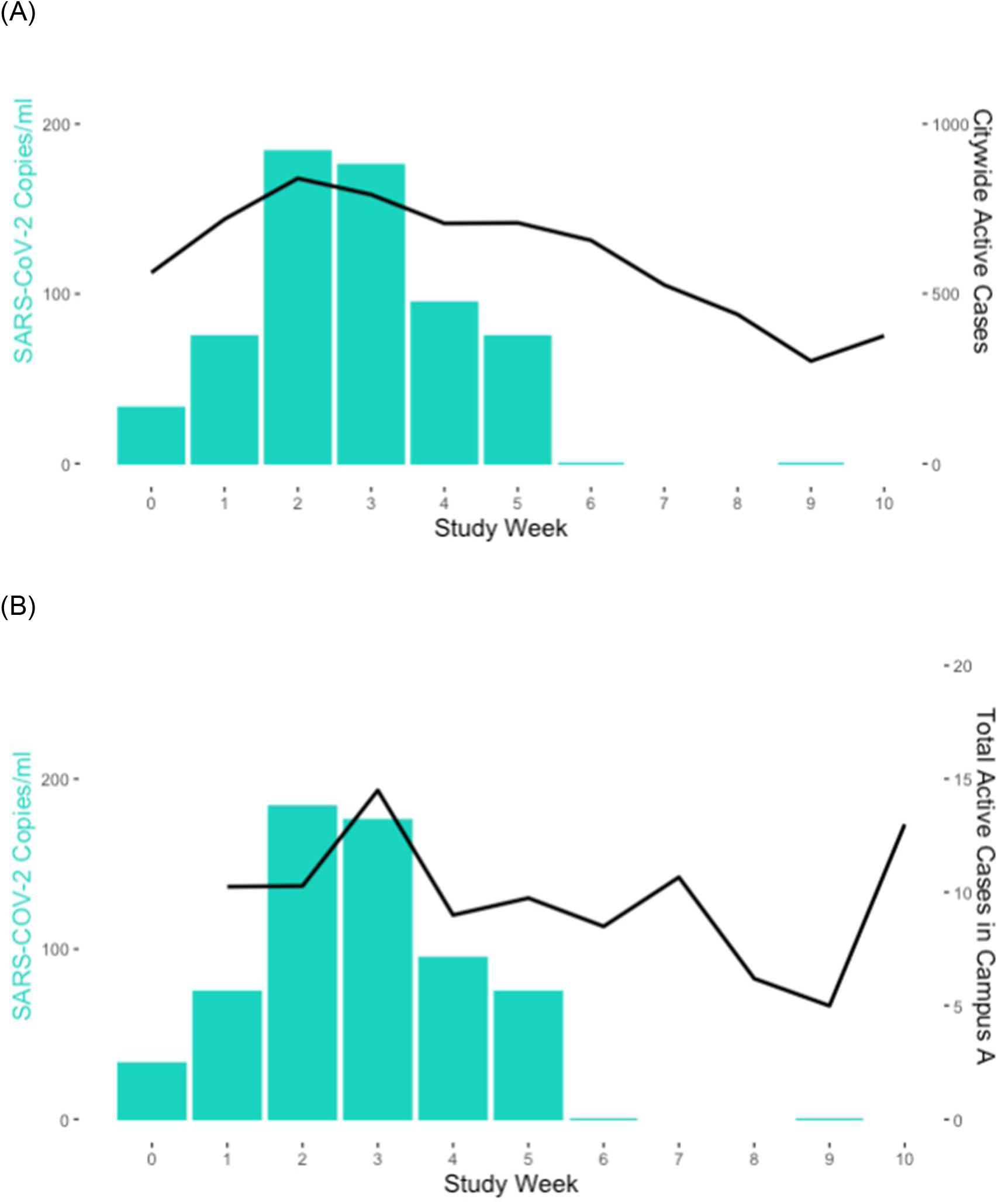
Campus A wastewater SAR-CoV-2 copies per ml (bars) and [A] mean citywide COVID-19 active cases (line) or [B] mean hospital COVID-19 active cases (line). There was no sample collection on weeks 7, 8 and 10.

We also evaluated the strength of association between (i) all built environment swab positivity and (ii) floor swab positivity with hospital case burden (Table 4). We found that floor swabs best described the hospital case burden, having a measure of association with the greatest precision, and the model with the lowest AIC (Table 4).

## DISCUSSION

In this prospective longitudinal study at two large tertiary care hospital campuses, we demonstrate that SARS-CoV-2 RNA can be readily identified from a variety of hospital surfaces as well as hospital wastewater. Moreover, both the relative frequency of isolation on surfaces and the viral titre in wastewater parallel the burden of infection within the hospital. Wards that specifically managed COVID-19 patients had consistently higher levels of detection compared to other areas, and frequency of detection paralleled case counts over time. For the single ward-based outbreak that occurred during the study period, there was detectable viral RNA at the involved location from the onset and until resolution.

Prior studies have demonstrated the amplification of SARS-CoV-2 RNA from the hospital environment^7^ including high touch surfaces outlined in this paper (*e*.*g*., elevator buttons, computer keyboards, alcohol hand pumps). Floors have been identified as a common location for identifying SARS-CoV-2 in hospitals, and our results reinforce this finding^7^. Moreover, we found that performing surveillance from floors provided the best marker for predicting hospital burden of infection.

To date, fomites have not been identified as a major contributor to transmission of SARS-CoV-2 in the hospital or other settings^5^. While our PCR-based approach for SARS-CoV-2 RNA detection cannot identify whether viral particles present in our samples were viable or not, our results do indicate that identifying any RNA from high touch surfaces is relatively rare, further supporting these as unlikely pathways for transmission in hospitals. Floors, however, were a common location for SARS-CoV-2 RNA in our study. It is likely they represent a sink for viral particles which originate from the respiratory droplets/aerosol from patients, health workers, or visitors, and it is unlikely that floors represent a reservoir for transmission^7^.

Interestingly, the non-COVID ward, Ward 3 on Campus A, that experienced the outbreak had frequent detection of SARS-CoV-2 during the outbreak period, with general absence before and after. This suggests that built environment sampling may offer a way to potentially identify occult outbreaks or transmission occurring within patients/visitors/healthcare workers or for monitoring outbreak status. The outbreak that occurred in the ED at Campus B did have a brief but non-sustained signal, and we believe this is related to the transient nature of patients (and flux of staff) within the ED. Patients are flowing from the ED itself to either discharge or transfer to other sites within the hospital, thus it is unlikely to sustain a patient-based outbreak within that area. The reason for higher detection of SARS-CoV-2 at Campus A is likely driven by more frequent detection at non-COVID risk unit sites. The reason for this is unclear and may relate to both patient and visitor populations.

Where built environment sampling can provide a resolved view of COVID-19 burden in the hospital, hospital wastewater can provide institution-wide surveillance with the possibility for anticipatory (predictive) signals. Qualitatively, wastewater appeared to anticipate burden by 1-2 weeks, and is on the order of magnitude reported in the literature^20^, though given the limited number of time points, we were not able to confirm this statistically. A growing number of studies have shown the potential benefit of wastewater monitoring in both regional as well as facility-based surveillance^20,21^, and our study links this surveillance as complementary to built environment screening. Combined approaches may ultimately be applicable to other respiratory viruses that cause significant institutional outbreaks, including influenza, though further validation is needed.

Our study has some limitations which have not already been noted. Firstly, our study was performed during a period when there was relatively low activity, with regional case counts ranging from approximately 2 to 7 cases per 100,000 population per week, and it is possible that in higher burden settings there could be saturation of ward-based built environment detection which could limit the correlation with burden or outbreaks. Secondly, outbreaks were relatively rare in our hospital, owing in part to the low regional burden, and this limits our ability to evaluate the ability of built environment screening to consistently detect outbreaks. Lastly, our approaches only detect viral RNA and not viable virus. However, our findings benefit from the generalizability of our approaches which use standard PCR techniques and could be easily and quickly performed in most settings worldwide.

In conclusion, we have demonstrated that the detection of SARS-CoV-2 from wastewater and built environment samples, in particular from floors, parallels hospital burden. Built environment sampling may offer a spatially-resolved method for surveillance of hospital COVID-19 cases which can be paired with wastewater monitoring for a comprehensive picture of facility-wide burden of infection. Further prospective studies are needed to evaluate this approach.

## Data Availability

The data that support the findings of this study are available from the corresponding author upon reasonable request.

## Acknowledgements

We would like to thank Dr. Kathryn Suh and Dr. Raphael Saginur for their support and advice during this study. We would also like to thank the City of Ottawa Wastewater Collection Branch, and in particular Benjamin Musyoka and Sandra Gay, for accomodating the collection of wastewater samples.

## Author’s contributions

All authors contributed to the design of the study. LX, DM, CN collected built environment samples for the study and performed data entry. RJK and AH processed samples for the study. DM and CN contributed to data analyses. All authors contributed to the writing of the manuscript.

## REFERENCES

1. Siemieniuk, R. A. C. et al.. Drug treatments for covid-19: living systematic review and network meta-analysis. BMJ 370, (2020).

2. Baden, L. R. et al.. Efficacy and Safety of the mRNA-1273 SARS-CoV-2 Vaccine. N. Engl. J. Med. 384, 403–416 (2021).

3. The Visual and Data Journalism Team. Covid map: Coronavirus cases, deaths, vaccinations by country. BBC News https://www.bbc.com/news/world-51235105 (2021).

4. Islam, M. S. et al.. Current knowledge of COVID-19 and infection prevention and control strategies in healthcare settings: A global analysis. Infect. Control Hosp. Epidemiol. 41, 1196–1206 (2020).

5. Mondelli, M. U., Colaneri, M., Seminari, E. M., Baldanti, F. & Bruno, R. Low risk of SARS-CoV-2 transmission by fomites in real-life conditions. Lancet Infect. Dis. (2020) doi:10.1016/S1473-3099(20)30678-2.

6. van Doremalen, N. et al.. Aerosol and Surface Stability of SARS-CoV-2 as Compared with SARS-CoV-1. N. Engl. J. Med. 382, 1564–1567 (2020).

7. Guo, Z.-D. et al.. Aerosol and Surface Distribution of Severe Acute Respiratory Syndrome Coronavirus 2 in Hospital Wards, Wuhan, China, 2020. Emerg. Infect. Dis. 26, 1583–1591 (2020).

8. Smith, D. R. M. et al.. Optimizing COVID-19 surveillance in long-term care facilities: a modelling study. BMC Med. 18, 1–16 (2020).

9. Westhaus, S. et al.. Detection of SARS-CoV-2 in raw and treated wastewater in Germany - Suitability for COVID-19 surveillance and potential transmission risks. Sci. Total Environ. 751, 141750 (2021).

10. Gonçalves, J. et al.. Detection of SARS-CoV-2 RNA in hospital wastewater from a low COVID-19 disease prevalence area. Sci. Total Environ. 755, (2021).

11. CDC. Labs. https://www.cdc.gov/coronavirus/2019-ncov/lab/rt-pcr-panel-primer-probes.html (2020).

12. Klymus, K. E. et al.. Reporting the limits of detection and quantification for environmental DNA assays. Environmental DNA 2, 271–282 (2020).

13. Public Health Ontario. https://www.publichealthontario.ca/-/media/documents/b/2020/bp-novel-respiratory-infections.pdf?la=en.

14. Ontario Ministry of Health. http://health.gov.on.ca/en/pro/programs/publichealth/coronavirus/docs/2019_reference_doc_symptoms.pdf.

15. Medema, G., Heijnen, L., Elsinga, G., Italiaander, R. & Brouwer, A. Presence of SARS-Coronavirus-2 RNA in sewage and correlation with reported COVID-19 prevalence in the early stage of the epidemic in the Netherlands. Environ. Sci. Technol. Lett. 7, 511–516 (2020).

16. Daily COVID-19 Dashboard. https://www.ottawapublichealth.ca/en/reports-research-and-statistics/daily-covid19-dashboard.aspx (2021).

17. Ontario Ministry of Health. http://www.health.gov.on.ca/en/pro/programs/publichealth/coronavirus/docs/2019_acute_care_guidance.pdf.

18. Gomila, R. Logistic or linear? Estimating causal effects of experimental treatments on binary outcomes using regression analysis. J. Exp. Psychol. Gen. (2020) doi:10.1037/xge0000920.

19. Parker, C. W. et al.. End-to-End Protocol for the Detection of SARS-CoV-2 from Built Environments. mSystems 5, (2020).

20. SARS-CoV-2 RNA in wastewater anticipated COVID-19 occurrence in a low prevalence area. Water Res. 181, 115942 (2020).

21. Sharif, S. et al.. Detection of SARs-CoV-2 in wastewater, using the existing environmental surveillance network: An epidemiological gateway to an early warning for COVID-19 in communities. medRxiv 2020.06.03.20121426 (2020).

